# Integrating large scale genetic and clinical information to predict cases of heart failure

**DOI:** 10.1101/2022.07.19.22277830

**Authors:** Kuan-Han H. Wu, Brooke N. Wolford, Xianshi Yu, Nicholas J. Douville, Michael R. Mathis, Sarah E. Graham, Global Biobank Meta-analysis Initiative (GBMI), Ida Surakka, Whitney E. Hornsby, Jiang Bian, Lili Zhao, Cristen J. Willer, Xu Shi

## Abstract

**Background:** Heart failure (HF) is a major cause of death globally. Prediction of HF risk and early initiation of treatment could mitigate disease progression.

**Objectives:** The study aimed to improve the prediction accuracy of HF by integrating genome-wide association studies (GWAS)- and electronic health records (EHR)-derived risk scores.

**Methods:** We previously performed the largest HF GWAS to date within the Global Biobank Meta-analysis Initiative to create a polygenic risk score (PRS). To extract clinical information from high-dimensional EHR data, we treated diagnosis codes as ‘words’ and leveraged natural language processing techniques to create a clinical risk score (ClinRS). Our method first learned code co-occurrence patterns and extracted 350 latent phenotypes (low-dimensional features) representing EHR codes, then used coefficients from regression of HF on the latent phenotypes in a training set as weights to calculate patient ClinRS in a validation set. Model performances were compared between baseline (age and sex) model and models with risk scores added: 1) PRS, 2) ClinRS, and 3) PRS+ClinRS. We further compared the proposed models with Atherosclerosis Risk in Communities (ARIC) heart failure risk score.

**Results:** Results showed that PRS and ClinRS were each able to predict HF outcomes significantly better than the baseline model, up to eight years prior to HF diagnosis. By including both PRS and ClinRS in the model, we achieved superior performance in predicting HF up to ten years prior to HF diagnosis, two years earlier than using a single risk predictor alone. Additionally, we found that ClinRS performed significantly better than ARIC model at one year prior to disease diagnosis.

**Conclusions:** We demonstrate the additive power of integrating GWAS- and EHR-derived risk scores to predict HF cases prior to diagnosis.

## INTRODUCTION

Heart failure (HF) affects an estimated 64 million patients worldwide with a growing burden anticipated as the population ages^1,2^. Echocardiographic screenings in the general population have revealed that up to half of the individuals living with heart failure may be undiagnosed, hampering earlier access to mortality-reducing treatments^3,4^. Applying risk prediction tools enables earlier identification of diseases, thereby shifting the trajectory of disease progression towards prevention. Additionally, gaining a deeper understanding of the key risk factors for heart failure could shed insight into the mechanisms of disease progression and guide therapeutic management. We sought to evaluate the predictive accuracy of a modern risk assessment tool that incorporates diverse clinical and genetic data compared to genetic or clinical prediction models alone^4–6^.

Clinical prediction tools, such as Framingham and atherosclerotic cardiovascular disease (ASCVD) risk score (also known as the Pooled Cohort Equation [PCE]), are commonly used to predict cardiovascular disease (CVD), which have been widely applied and updated over time to include a variety of demographic, laboratory, hemodynamic, and medical details^7–11^. Researchers have established risk scores to predict the risk of developing heart failure^12^. However, due to the heterogeneous nature of heart failure, it is difficult to fully capture the risk solely on clinical data, which could lead to incomplete assessment as it overlooks genetic information that contributes to the remaining risk ^13–15^. Novel risk scores incorporating both diverse clinical data and well-powered genetic data are needed for a more precise prediction of heart failure risk.

Genome-wide polygenic risk scores (PRS) estimate an individual’s cumulative genetic risk for a given disease as a weighted sum of genetic effects estimated from GWAS for thousands to millions of genetic variants. Multiple studies have demonstrated that incorporating a PRS into disease risk prediction can enhance prediction accuracy and further improve early prevention^6,16^. Multiple efforts have been made to summarize genetic and clinical information for early identification of high-risk patients, but integrating high-dimensional genome-wide association study (GWAS) and electronic health record (EHR) into heart failure prediction models has yet to be assessed^17–19^.

We explore approaches to enhance the prediction of heart failure events leveraging both genetic and clinical data. See Figure 1 for the central illustration of this study, which integrates recent insights on the genetic underpinning of heart failure with a novel EHR-based clinical scoring system, referred to as the clinical risk score (ClinRS), to predict heart failure. The PRS was powered by the largest heart failure GWAS^20^ to date, while the clinical risk assessment borrowed natural language processing (NLP) techniques to capture co-occurrence patterns of medical events within the structured EHR data. From the proposed approaches above, we summarized 907,272 genetic variants into a PRS and 29,346 medical diagnosis codes into a ClinRS. We hypothesized that the additive power of integrating PRS and ClinRS would result in the most powerful heart failure prediction model.

**Figure 1.**
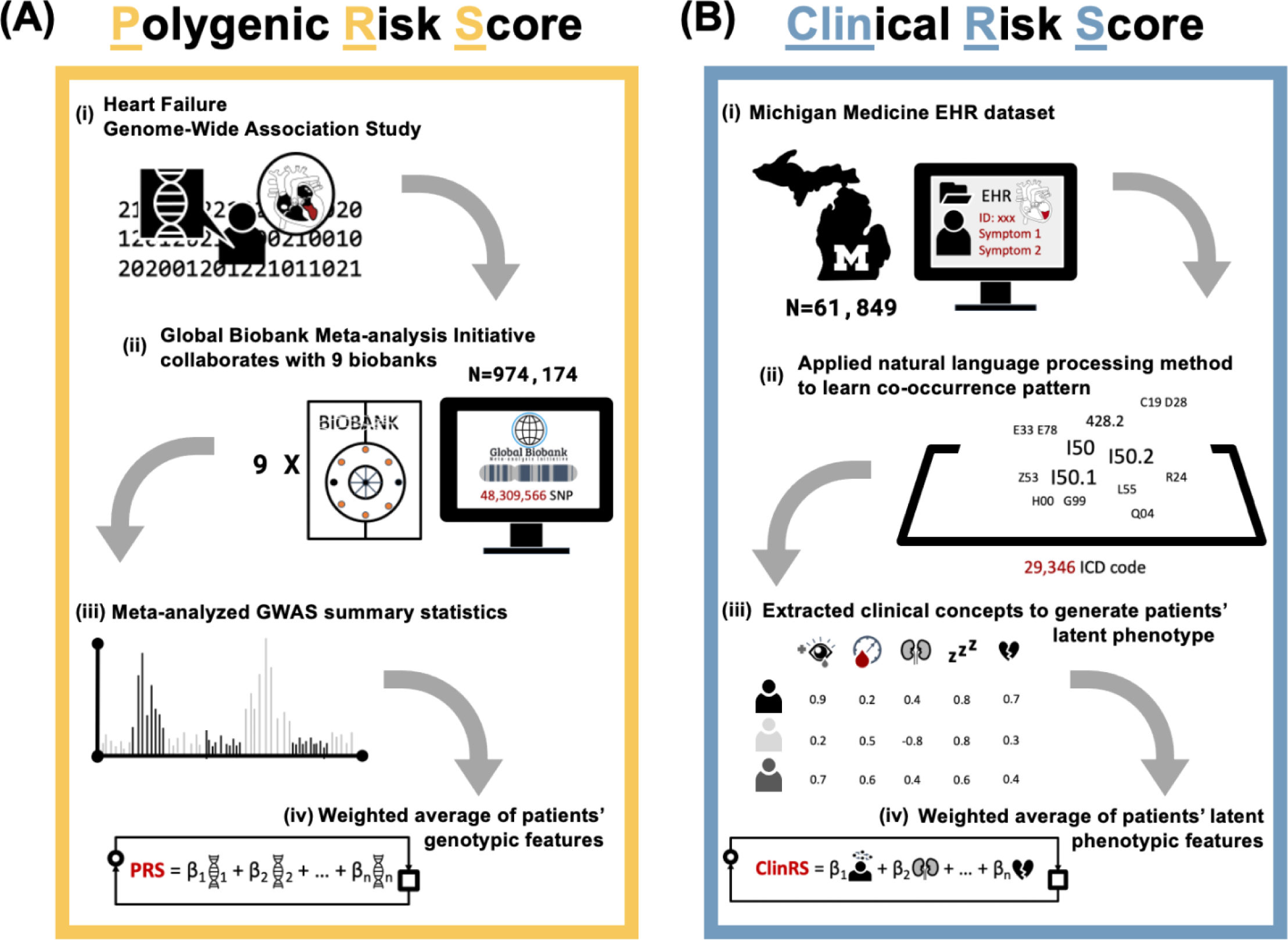
This study leveraged genetic and clinical data to improve heart failure cases prediction. **(A)** Polygenic risk score (PRS) was generated (i) using the largest heart failure genome-wide association study (GWAS). (ii) Heart failure GWAS were conducted in nine biobanks with total of 974,174 individuals and meta-analyzed by Global Biobank Meta-analysis Initiative. (iii) The meta-analyzed summary statistics were further used to calculate individuals’ PRS, (iv) a weighted average of the genotypic risk. **(B)** Clinical risk score (ClinRS) was created using novel natural language processing method to extract clinical concepts of each EHR code and summarize patients’ phenotypic risk. (i) Michigan Medicine EHR data of 61,894 patients with 29,346 unique codes were used to (ii) learn code co-occurrence patterns and the information were further used to (iii) generate patients’ latent phenotype. (iv) Next, we regressed heart failure outcome on patients’ latent phenotypes to derive the weights of ClinRS.

## METHODS

To generate the most statistically powered genetic predictor, we meta-analyzed multiple biobank datasets within the Global Biobank Meta-analysis Initiative (GBMI) consortium to generate a heart failure GWAS^20,21^. The GBMI consortium aims to enhance GWAS power and improve disease risk prediction through international collaboration among biobanks across the world and making all GWAS summary statistics open-access for researchers. The case count of the heart failure GWAS from GBMI is the largest-to-date and the PRS generated from GBMI meta-analysis GWAS is showed to have higher accuracy in predicting future heart failure events^20^.

To extract clinical information from EHR, we developed novel machine learning methods to efficiently summarize large-scale structured EHR data into a ClinRS for heart failure. We treated medical diagnosis codes (i.e., International Classification of Diseases [ICD] code) as ‘words’ in human language and adapted NLP methods to capture the co-occurrence pattern among codes in the high-dimensional medical records. The co-occurred relationship among codes was then utilized to extract independent information and converted into low-dimensional numeric vectors resembling the context and semantics of medical events. The University of Michigan’s Institutional Review Board approved these protocols (HUM00128472 and HUM00143523).

Three cohorts of Michigan Medicine (MM) patients were used in this study: 1) Primary Care Provider cohort (MM-PCP; N=61,849), 2) Heart Failure cohort (MM-HF; N=53,272), and 3) Michigan Genomics Initiative cohort (MM-MGI; N=60,215). The data were recorded between 2000 to 2022 in the Michigan Medicine EHR system, which includes both ICD-9 and ICD-10 diagnosis codes. See Supplementary methods section, “Michigan Medicine EHR system and biobank,” as well as Supplementary Figure 1 for detailed description of the MM-PCP, MM-MGI, and MM-HF cohorts.

### Polygenic Risk Score (PRS)

The polygenic risk score was derived from the heart failure GWAS conducted by the Global Biobank Meta-analysis Initiative. GBMI is a global collaboration network of 23 biobanks, across 4 continents and with over 2.2 million participants (as of April 2022)^21^. The summary statistics from nine of the GBMI heart failure contributing cohorts (BioMe, BioVU, Estonian Biobank, FinnGen, HUNT, Lifelines, Partners Biobank, UCLA Precision Health BioBank, and UK Biobank) were meta-analyzed resulting in 974,174 individuals of European ancestry in the combined GWAS. These nine biobanks contributed a total of 51,274 heart failure cases and 922,900 healthy controls, defined by phenome-wide association study code (phecode)^22^ 428.2 (heart failure, not otherwise specified)^22,23^. The GBMI heart failure study has the highest heart failure case number in a published GWAS study to date. Advanced genotyping imputation reference panels were used in the participating cohorts, resulting estimates of variation at additional genetic variants. These advancements in the GBMI heart failure GWAS improved the statistical power to more precisely identify the genetic risk associated with the outcome^24,25^. In this study, we used the GBMI European-ancestry meta-analysis GWAS to generate a heart failure PRS, which is the current best performing heart failure PRS for European American individuals^20^. See Figure 1, part A, illustrating the PRS calculation.

The weights used to create PRS were calculated with PRS-CS^26^, using European individuals from the 1000 Genome and UK Biobank combined cohort as the LD reference panel^27,28^. The meta-analyzed heart failure GWAS summary statistics from GBMI used in this study was independent from the validation set used in the analysis to compare the effect contribution between genetic and clinical information for predicting heart failure. PRS were calculated for individuals in the MM-MGI cohort. Possible population substructure was controlled by regressing the raw PRS on the top 10 principal components (PC) derived from the patient’s genotype file. The resulting residuals were inverse normalized to transform the final PRS score into a standard normal distribution.

### Clinical Risk Score (ClinRS)

To extract information from high-dimensional EHR data, we developed a novel clinical risk score, ClinRS, to summarize a patient’s longitudinal medical records into one single risk score via NLP techniques. First, we treated 29,346 EHR diagnosis ICD codes as ‘words’ and concatenated all codes documented in a patient’s whole medical history into an ‘article’ using the MM-PCP cohort. After we created the article from all patients, we adapted a NLP technique to obtain numeric vector representations that captured the semantic meaning and context of medical codes^29–31^. These vector representations were subsequently validated to be clinically meaningful, in the sense that it captured the concept of each code and showed high concordance with expert manually curated phenotypic grouping labels^22^. We refer to these representations as medical code embeddings hereon. See Supplementary methods section, “calculate clinical risk score – extraction of medical code embeddings using NLP,” for detailed description of creating medical code embeddings.

We leveraged the medical code embeddings to create ClinRS which is a linear combination of i) ClinRS weights from the ClinRS weights derivation set (MM-HF, excluding MM-MGI) and ii) patient latent phenotypes in the model validation set (intersection of MM-MGI and MM-HF). To create ClinRS weights, we first generated patient-level latent phenotypes that combines code embeddings and patients’ code utilization. See Supplementary methods section, “calculate clinical risk score – calculation of patient-level latent phenotypes,” for detailed explanation. We then regressed heart failure outcome on latent phenotypes, and the regression coefficients were utilized as weights for calculation of the ClinRS. See Supplementary methods section, “calculate clinical risk score – supervised training for ClinRS using LASSO,” for detailed explanation. Lastly, the trained ClinRS weights were combined with patient-level latent phenotypes in the validation set to create the final ClinRS. See Supplementary methods section, “calculate clinical risk score – calculate ClinRS for patients in model validation set,” for detailed explanation. With these steps, we successfully reduced the data dimension from 29,346 unique ICD codes to 350 latent phenotypes then to a single risk score. See Figure 1, part B, and Supplementary methods section, “calculate clinical risk score,” for detailed description of ClinRS curation.

### Statistical analysis

We conducted analyses within cohorts of 20,279 individuals in the model validation set (intersection of MM-MGI and MM-HF) with at least one year of medical history prior to a heart failure diagnosis in the Michigan Medicine health system (Supplementary Figure 1). Ten different datasets with time point cutoffs, one year apart from one year to ten years prior to disease diagnosis, were applied to the analysis. Individuals with no medical history prior to the time point cutoff were removed from the corresponding analysis.

We fit four logistic regression models to predict whether patients have a heart failure diagnosis, and further evaluated the accuracy among models with different risk predictor(s) for all ten time points, one to ten years prior to disease diagnosis. The baseline model included patients’ demographic information (age at diagnosis and sex), and three additional models with the risk score added: i) PRS, ii) ClinRS, and iii) PRS+ClinRS were created to compare the improvement in model accuracy from the baseline model. In the PRS and PRS+ClinRS models, the top ten PCs derived from patients’ genotype data were adjusted to account for the population structure^32^.

We further compared model performance with existing HF risk score, from Atherosclerosis Risk in Communities (ARIC) study^12^. The components of ARIC HF risk score include demographic information, vital signs, smoking history, medication history, disease diagnosis, and biomarker (N-terminal pro-B-type natriuretic peptide [NT-proBNP]). Two versions of ARIC HF risk score were published: 1) model without NT-proBNP and 2) model included NT-proBNP as predictor. In this study, we were only able to implement the model without incorporating NT-proBNP, as most of the patients do not have this biomarker available in our dataset. Missing information were imputed as zero for the predictors in ARIC risk score calculation to keep the sample size in testing set consistent across different risk score models.

Model performances were compared using 10-fold cross validated Area Under the Receiver Operating Characteristics (AUC). The analysis was performed using European ancestry samples only.

## RESULTS

In this paper, we utilized three independent datasets (Supplementary Figure 1) at Michigan Medicine to achieve two main goals: 1) obtain medical code embeddings using NLP techniques in EHR data and 2) improve heart failure prediction using PRS and ClinRS. First, we used the MM-PCP cohort with a total of 61,849 individuals and 159,273,800 diagnosis coding records from 2000 to 2022 to learn the medical code co-occurrence patterns and to extract medical code embeddings representing the clinical meaning of each code. The medical code embeddings trained from MM-PCP were validated using phenotype grouping labels to evaluate whether vector representations derived from an unsupervised NLP method are clustered in similar ways compared to expert manually curated code grouping (Supplementary Figure 2).

Next, we built two risk scores, PRS and ClinRS, in the model validation set (intersection of MM-MGI and MM-HF) to predict future heart failure cases. The PRS was calculated using heart failure GWAS summary statistics, meta-analyzed from nine biobanks in GBMI (independent from Michigan Medicine)^20^. We chose the European ancestry GWAS summary statistics (51,274 cases and 922,900 controls) as the base of our PRS to match the European ancestry of MM cohorts due to its superior performance in the European ancestry individuals in the original publication. From these summary statistics, a total of 907,272 genetic variants were integrated into a polygenic risk score.

The ClinRS calculation required two steps: i) create patient-level latent phenotypes and ii) derive weights (effect sizes) to calculate ClinRS. We generated medical code embeddings for 29,346 medical codes from MM-PCP, and then used the medical code embeddings to create 350 latent phenotypes for each patient in MM-HF. To derive weights for the ClinRS, we regressed heart failure outcome on latent phenotypes in ClinRS weights derivation set (MM-HF, excluding MM-MGI) and extracted the effect sizes as ClinRS weights. The ClinRS weights derivation set had a heart failure incidence of 330 out of 7,120 patients (4.6%) whereas in the model validation set we observed 576 (2.8%) heart failure cases out of 20,279 patients (Supplementary Figure 1). Our method integrated 29,346 medical diagnosis codes into a single clinical risk score (ClinRS). We further calculated ARIC heart failure risk score to compare the prediction performance of the proposed model with existing risk score. Details of ARIC HF risk score was described in statistical analysis section.

### NLP extracted medical code embeddings are clinically meaningful

First, we validated whether the medical code embeddings generated in MM-PCP cohort were clinically meaningful and suitable for generating a ClinRS, and whether the embeddings could capture the information hidden in the complex EHR dataset. We used the cosine distance between a pair of codes to classify whether a code pair shared the same expert manually curated phenotypic grouping, phecode, (i.e., have similar clinical concept) and calculated the concept-AUC. Concept-AUC is the AUC for identifying code pairs in the same phecode group, which was used to aid grid search for optimal NLP derived medical code embeddings based on existing clinical concept ontology. See detailed explanation in Supplementary method section, “evaluation of NLP derived medical code embeddings and parameter tuning.”

We found that diagnosis codes recorded on the same day provided the most information about code relationships (see details of the results in Supplementary result section, “NLP extracted medical code embeddings are clinically meaningful”). One possible explanation could be that diagnostic codes were often all billed on the same day, e.g., on the last day of hospitalization. Additionally, expanding the time window for code co-occurrence calculation could potentially introduce noise since diagnosis code recorded on different days may not be related to the same medical event.

We also found that the concept-AUC plateaued with up to embedding dimensions of 300 to 500, depending on the time-window. This finding is similar to previous reports^33–37^. We ultimately used medical code embeddings trained from co-occurrence patterns within the same day and with a dimension of 350, which yielded a concept-AUC of 0.78 (Supplementary Figure 2). This result supports that the medical code embeddings derived via unsupervised learning were clinically meaningful and validated by expert manually curated phenotypic grouping. The medical code embeddings corresponding to the above chosen tuning parameters were further used to calculate patient-level latent phenotype in this analysis.

In addition to numerically evaluating the semantic resemblance of vector representations using concept-AUCs, we further assessed the semantic relationship graphically using a heatmap of the cosine similarity scores (Supplementary Figure 3). In this study, we used ICD-9 Second Chapter (140-239): Neoplasms as an example to discern how the similarity patterns were formulated among each cancer code. Cancer codes were selected to demonstrate the similarity patterns of code pairs due to its distinct organ system specific sub-chapter within the cancer codes. For example, codes from cancer of digestive organs (ICD: 150-159) and cancer of respiratory organs (ICD: 160-165) are both cancer codes, but for different organs and were therefore expected to have different patterns and concepts.

As anticipated, we observed that the same ICD-9 diagnosis codes and/or nearby codes (off-diagonal line in Supplementary Figure 3) had higher cosine values between their embeddings, indicated by the darker color on the off-diagonal line and the band surrounding it. Furthermore, clear distinctions crossing different sub-chapters were found. These results suggest the contextual representations were clinically meaningful since related types of cancers from the same organ system had more similar context and patterns of co-occurred comorbidities, treatments, or procedures. Conversely, lower cosine scores were found in code pairs between different sub-chapters of cancer diagnosis ICD-9 codes.

### PRS and ClinRS each predict heart failure cases up to eight years in advance

We assessed the accuracy of using genetic and clinical information, individually, in predicting future heart failure. Our evaluation metric was 10-fold cross-validated AUC. We analyzed the performance of each risk score built from ten different time intervals, ranging from one year to ten years prior to the diagnosis of heart failure. To simplify, we refer to these ten intervals as ten time points. Sample size in each time interval decreased from one year to ten years prior to disease diagnosis, ranging from 20,279 (576 cases) to 10,391 (332 cases) participants, respectively (Supplementary Table 1). We thus had sufficient power to fit an accurate model given that the number of predictors, including demographic information, two risk scores, and PCs derived from genotype data, was less than fifteen.

We summarized the AUCs of the four models (baseline, PRS, ClinRS, and PRS+ClinRS model) and ARIC HF risk score each built from ten time points in Figure 2. Our results showed that both PRS and ClinRS models performed significantly better than the baseline model (which only included age and sex) up to eight years prior to heart failure diagnosis. The results were statistically significant, as determined by non-overlapping 95% confidence intervals (CI). This indicates that each risk score can individually predict heart failure diagnosis better than baseline. The highest AUC was observed in the ClinRS model (0.85 [95% CI: 0.83-0.87]) one year prior to diagnosis, followed by the PRS model (0.76 [0.74-0.83]), which were significantly higher compared to the baseline model with AUC of 0.70 (0.68-0.72). Overall, ARIC performed similar to ClinRS, both showed significantly better performance up to eight years prior to HF diagnosis, compared to baseline model. Additionally, we observed that at one year prior to HF diagnosis, ClinRS had significantly better performance on predicting HF cases, compared to ARIC. See Supplementary Table 1 for the specific AUC values across all ten time points and four different models. As expected, the benefits of risk scores derived from clinical information, ClinRS and ARIC, prediction were attenuated by censoring EHR data with increasing time thresholds prior to the event and decreasing sample size. However, better performance in ClinRS and ARIC were still observed until eight years prior to the disease diagnosis. On the other hand, model performances, using genetic information, were stable across all time points, which yielded significantly higher performance than baseline model from one year to eight years prior to the disease diagnosis. For example, in a cohort with at least eight years of medical history within Michigan Medicine, the PRS and ClinRS models yielded an AUC of 0.76 (0.74-0.78) and 0.77 (0.74-0.79), respectively, significantly higher compared to the baseline model with AUC of 0.71 (0.68-0.73).

**Figure 2.**
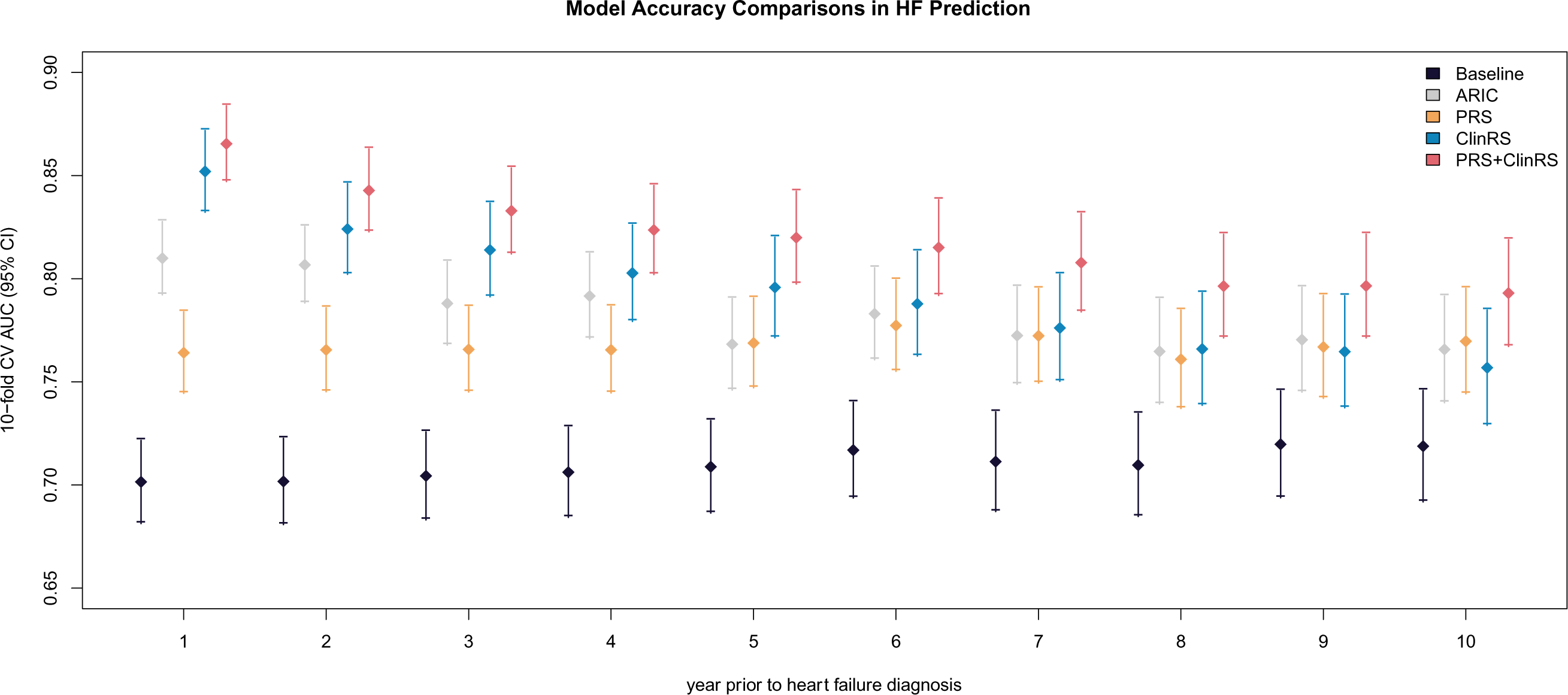
Forest plot comparing models’ accuracy of predicting heart failure at one to ten years prior to disease diagnosis. Five models were compared with each time point: baseline (age and sex), ARIC (Atheroscierosis Risk in Comunities heart failure risk score), PRS (polygenic risk score), ClinRS (clinical risk score), and PRS+ClinRS. Numbers at the bottom of the plot indicate the sample size for each time point. Results showed that ARIC, PRS, and ClinRS, separately, can predict heart failure outcomes eight years in advance, and adding both risk predictors in the model can predict disease ten years in advance. Additionally, ClinRS performed significantly better than ARIC in predicting heart failure event in one year.

In models given data from nine years prior to disease diagnosis, no significant difference was observed among, PRS (AUC: 0.77 [0.74-0.79]), ClinRS (AUC: 0.76 [0.74-0.79]), ARIC (AUC: 0.77 [0.75-0.80]), and baseline (AUC: 0.72 [0.69-0.75]) models. This lack of significant difference between PRS, ClinRS, ARIC, and the baseline model from data nine years before the diagnosis could potentially be attributed to the smaller sample size. The limited information provided by the EHR data nine and ten years prior to the disease diagnosis may have also contributed to the lack of significantly increased prediction, as it may have not provided enough information for complex prediction tasks.

### Integrating PRS and ClinRS enhances heart failure prediction

In addition to evaluating the risk score separately, we further studied the additive power of including both risk scores together in the heart failure prediction model. Consistently across all ten time points, the highest accuracy was found in the PRS+ClinRS model. See Figure 2 and Supplementary Table 1. Significantly higher AUC was continuously found in the PRS+ClinRS model even at ten years prior to disease diagnosis with an AUC of 0.79 (0.77-0.82), compared to baseline model (AUC: 0.72 [0.69-0.75]). Compared to the single risk predictor models predicted heart failure eight years prior to disease diagnosis, the model including both predictors predicted disease two years earlier than using either single risk predictor alone.

As expected, we observed that the prediction accuracy of the PRS+ClinRS model outperformed single risk score models throughout the entire one to ten years time horizons. In Supplementary Figure 4, we showed that by using both clinical and genetic risk scores to predict which individuals have a high risk of future heart failure, the combined score identified the highest proportion (28%) of individuals who had heart failure.

### ClinRS insights

We dissected the composition of ClinRS for heart failure prediction and further studied the risk and protective factors associated with disease outcome in Supplementary Figure 5. In Supplementary Figure 5, we showed the ClinRS weights of risk and protective factors contributing to the heart failure outcome. The diagnoses prioritized in the ClinRS score can generally be classified by 1) organ system (cardiac versus non-cardiac) and 2) etiology (potential causal mechanism, associated comorbidity, or unclear link). As expected, seven out of the top ten risk factor for heart failure in ClinRS were cardiac diagnoses, exhibiting potential causal mechanisms; for example, ICD-9 codes associated with acute myocardial infarction (Supplementary Table 2). Additional potential causal diagnoses for HF diagnoses including: i) stenosis, mitral and aortic valves (ICD: 396.0), ii) acute myocarditis (ICD: 422.0), and iii) defect, acquired cardiac septal (ICD: 429.71) were highly prioritized by the ClinRS algorithm. Also, ClinRS incorporates many associated-cardiac diagnoses including i) malfunction, cardiac pacemaker (ICD: 996.01) and ii) mechanical complication of automatic implantable cardiac defibrillator (ICD: 996.04). These codes are likely to co-occur in patients with heart failure, but may have limited utility in predicting new or previously undiagnosed cases; although it is noteworthy that all diagnoses included in ClinRS were documented prior to the heart failure diagnosis. Diagnoses identified by ClinRS including: i) Marfan syndrome (ICD: 759.82, 754.82)^38^, ii) alcohol abuse (ICD: 303.01, 790.3, 980.0)^39^, and iii) viral infection (ICD: 74.8)^40^ may reflect non-cardiac, causal mechanisms of heart failure pathogenesis. Notably, non-cardiac-related diagnoses, unclear link with a protective effective against heart failure, in the ClinRS score included a cluster of pregnancy-related conditions (ICD: 765.14, 765.25, 656.43, 678, etc) and another cluster of ophthalmologic diagnoses (ICD: 371.03, 370.03, 370.63, 374.23, 370.35, etc). No causal or mechanistic relationship should be inferred. This correlation likely results from the lower-risk baseline population (childbearing females) for pregnancy related-conditions and more focused, clinical ophthalmologic assessment being less likely to diagnose heart failure, for the ophthalmologic-conditions.

## DISCUSSION

This study sought to improve the accuracy of heart failure prediction by integrating high-dimensional genetic data with clinical information to further heart failure prevention initiatives. Genetic risk was summarized by a PRS, calculated from the largest-to-date heart failure GWAS^20^, and clinical risk was summarized by a ClinRS, a novel EHR-based risk score. The combined PRS and ClinRS score prediction model identified patients with a high risk of heart failure a decade in advance of the disease diagnosis (Figure 2 and Supplementary Table 1). Specifically, the PRS+ClinRS prediction model showed a significantly higher AUC at ten years prior to heart failure diagnosis with AUC of 0.79 (0.77-0.82) compared to the baseline model with AUC of 0.72 (0.69-0.74). In contrast, models relying on a single risk score can only identify heart failure cases eight years in advance. However, by integrating genetic and clinical information we identify heart failure cases two years earlier. These findings reveal the power of integrating PRS and ClinRS to enhance disease prediction and the potential to inform heart failure prevention efforts. More broadly, this study highlights the methods and opportunity to curate ClinRS for other complex diseases and integrate with PRS to improve disease prediction accuracy.

### Advances in comprehensively utilizing longitudinal and high-dimensional EHR data

The critical challenges of incorporating EHR data are its high dimensionality and longitudinal nature. We successfully developed a risk score summarizing the clinical information despite the complexity of EHR data and validated its utility in an independent dataset from an EHR-linked biobank cohort. This study treated structured EHR diagnosis codes as human language and converted the diagnosis code into articles. This enabled learning the coding patterns for patient records with any dimensionality and longitudinal history. By focusing on co-occurrence patterns of medical codes within a specified time window, we were able to utilize data from all individuals regardless of the length of healthcare utilization. In addition, by applying NLP to transform codes to medical code embeddings, we successfully reduced the high-dimensional EHR dataset into low-dimensional features. The results present an avenue to incorporate other domains of structured EHR datasets, such as medical procedures and laboratory tests, to create a clinical risk score that could more comprehensively capture the risk of having the disease.

### An integrated model (PRS + ClinRS) enables improved prediction of heart failure

We previously developed a heart failure GWAS with the largest number of cases to date to build heart failure risk prediction models^20^. We successfully reduced high-dimensional GWAS into a single predictor – PRS. Furthermore, we implemented adapted NLP techniques to capture latent phenotypes in EHR data and summarized them into a new predictor – ClinRS. The analysis results showed that both the PRS and ClinRS were significantly better predictors of heart failure compared to baseline demographic information alone. Additionally, including both PRS and ClinRS together into an additive prediction model yielded superior accuracy for predicting future heart failure outcomes. This result demonstrated the additive predictive power of leveraging genetic and clinical information in risk prediction.

In alignment with our findings, Mujwara et al., used CAD-PRS to reclassify high genetic risk patients from patients in the borderline or intermediate of PCE clinical risk pool^18^. Their work showed that using the combined PCE and CAD-PRS approach risk screening methods to initiate early preventive treatment could potentially avert 50 ASCVD events over 10 years per 10,000 individuals screened and lead to substantial cost saving per averted event. It is promising that we have the potential to achieve more accurate prediction by using PRS and ClinRS together into prediction models. Such strategies could then inform guidelines for patient care to aid in earlier initiation of prevention treatment.

### Medical code embeddings filled in missing information/incomplete EHR history

We strengthened the evidence that leveraging genetic and clinical information improves precision health by performing a sensitivity analysis with all circulatory system diagnosis codes removed (see Supplementary results section, “sensitivity analysis on removing circulatory system diagnosis code”). Despite partially missing clinical information from the EHR system, we were still able to reach high prediction accuracy one decade prior to disease diagnosis by incorporating ClinRS, without circulatory system diagnosis code, and PRS in the full model (Supplementary Figure 6 and Supplementary Table 1). This analysis also indicated the potential benefit for patients with short medical history within the same healthcare system, missing information and/or unrecorded diagnosis would be able to reveal from the incomplete health records using pre-trained medical code embedding^41^.

Furthermore, the prediction ability of applying medical code embeddings to fill in the missing information from incomplete EHR records showed that ClinRS could be a more scalable approach compared to traditional risk score calculation. To calculate traditional risk score with multiple clinical information and biomarkers in large population, it required tremendous resources to collect patients’ data and it could lead to underdiagnosis for population with less access to healthcare. While using pretrained medical code embeddings, we would be able to borrow information from other patients with similar medical condition patterns to predict risk for patients with less healthcare visit or missing medical records.

### ClinRS is superior to traditional risk score in prediction and calculation

The ARIC heart failure risk score was curated from experts by pre-selecting risk predictors to fit into the prediction model^12^. The predictors used in calculating ARIC HF risk score required intensive labors to survey patients’ health outcome, measure biomarkers to identify diabetes patients, adjudicate coronary heart disease status from electrocardiography by clinician, and so on. In a non-cohort study setting, often time patients will not have the full sets of predictors available in their health records, which could lead to underperformance of the disease prediction. Hence, it is important to develop a method that could perform equally well with or without missing information.

Here, we demonstrated using existing large scale EHR-linked biobank to create standardizable and scalable risk predictors. By applying NLP method to extract the co-occurrence patterns among patients’ healthcare utilization, we successfully built a risk score, ClinRS, with high performance in predicting HF cases. Moreover, ClinRS, used unsupervised approach to select predictors, yielded a significantly higher prediction accuracy at one year prior to HF diagnosis, compared to ARIC, pre-selected risk predictors by experts.

### Study limitations

Heart failure is known to have separate subtypes with distinct treatments and phenotypic symptoms caused by different mechanisms, environmental exposures, or genetic risk factors^20^. In the future, ClinRS for heart failure subtypes should be developed and validated in cohorts with larger sample sizes. Moreover, the curation of ClinRS and utilization of integrating genetic and clinical information for disease risk prediction needs to be benchmarked in other complex diseases.

This study solely utilized the diagnosis information derived from EHR data, however, leveraging other domains of structured and unstructured EHR data (e.g., procedure, medication, clinical notes, etc.) to assist disease prediction is needed to fully understand the additive power of integrating genetic and clinical data.

Furthermore, the limitation of EHR-based study also includes the low transferability across different healthcare systems, due to the heterogeneity of EHR data. Methodology in language models could potentially be borrowed to improve transferability of medical code embeddings and the derived latent phenotypes. Applying transfer learning techniques could also produce a more generalizable ClinRS to be applied across different healthcare systems.

Lastly, due to the limited sample size of people of diverse ancestral background, we were only able to perform analysis in European American cohort in the MGI biobank. In the future, models validated in diverse population in needed.

### Conclusion

In conclusion, the amalgamation of GWAS- and EHR-derived risk scores predicted heart failure cases 10-years prior to diagnosis. These findings highlight how application of natural language processing to complex datasets such as medical records and incorporation of genetic information may enhance the identification of patients with a higher susceptibility to heart failure. Application of this approach at scale may enable physicians to implement preventive measures at a much earlier stage, potentially up to eight years before a would-be diagnosis, which may prevent the onset of overt heart failure.

## Data Availability

All data produced in the present study are available upon reasonable request to the authors

## Acknowledgements

We would like to express our gratitude to all contributors to GBMI and the biobank participants who provided their data for biomedical research. The authors acknowledge the participants, recruitment teams and project managers of the Global Biobank Meta-analysis Initiative for providing data aggregation, management, and distribution services in support of the research reported in this publication (particularly Sinéad Chapman and Bethany Klunder). The authors would like to acknowledge Da-Wei Lin (University of Michigan) for the help with central illustration in Figure 1.

This work was supported by the National Institutes of Health grants T32-GM070449 (KHW) from the National Institute of General Medical Sciences, K08-DK131346 (NJD) from the National Institute of Diabetes and Digestive and Kidney Diseases, K01-HL141701 (MRM) from the National Heart, Lung, and Blood Institute, R35-HL135824 (CJW) from the National Heart, Lung, and Blood Institute, and R01-GM139926 (XS) from the National Institute of General Medical Sciences.

