## Supplementary Document for "Integrating large scale genetic and clinical information to predict cases of heart failure"

**Supplementary Materials for "Integrating large scale genetic and clinical information to predict cases of heart failure"**

### Author information

Kuan-Han H. Wu, PhD[1], Brooke N. Wolford, PhD[2], Xianshi Yu, PhD[3], Nicholas J. Douville, MD, PhD[1,4,5], Michael R. Mathis, MD[1,4,5], Sarah E. Graham, PhD[6], Global Biobank Meta-analysis Initiative (GBMI), Ida Surakka, PhD[6], Whitney E. Hornsby, PhD[6], Jiang Bian, PhD[7], Lili Zhao, PhD[8], Cristen J. Willer, PhD[1,6,9*], and Xu Shi, PhD[4*]

### Affiliations

[1] Department of Computational Medicine and Bioinformatics, University of Michigan, Ann Arbor, Michigan, USA
[2] K.G. Jebsen Centre for Genetic Epidemiology, Department of Public Health and Nursing, Norwegian University of Science and Technology (NTNU), Trondheim, Norway
[3] Department of Biostatistics, University of Michigan, Ann Arbor, Michigan, USA
[4] Department of Anesthesiology, Michigan Medicine, Ann Arbor, Michigan, USA
[5] Institute of Healthcare Policy & Innovation, University of Michigan, Ann Arbor, Michigan, USA
[6] Department of Internal Medicine, Michigan Medicine, Ann Arbor, Michigan, USA
[7] Health Outcomes & Biomedical Informatics, College of Medicine, University of Florida
[8] Beaumont Health
[9] Department of Human Genetics, University of Michigan, Ann Arbor, Michigan, USA
[*] Senior author

### Correspondence

Xu Shi, PhD

### METHODS

#### Michigan Medicine EHR system and biobank

Three cohorts of Michigan Medicine (MM) patients were used in this study: 1) Primary Care Provider cohort (MM-PCP; N=61,849), 2) Heart Failure cohort (MM-HF; N=53,272), and 3) Michigan Genomics Initiative cohort (MM-MGI; N=60,215) (Supplementary Figure 1). All individuals in the three cohorts underwent at least one surgical procedure within the MM healthcare system. The data were recorded between 2000 to 2022 in the Michigan Medicine EHR system, which includes both ICD-9 and ICD-10 diagnosis codes.

The MM-PCP cohort includes patients i) with primary care providers within Michigan Medicine, ii) who had received an anesthetic, iii) whose most recent visit was in 2018 or later, and iv) who had at least five years of medical encounter history (difference between last and first encounter year greater or equal to five) within Michigan Medicine. Exclusion criteria for this cohort includes patients i) recruited in the Michigan Genomics Initiative and ii) predefined in the Heart Failure cohort to ensure that no samples overlap with datasets used to validate the clinical predictor.

The MM-HF cohort was defined by a previously validated heart failure phenotyping algorithm[1]. The phenotyping algorithm incorporated ICD diagnosis codes, medication history, cardiac imaging, and clinical notes in the form of free text to assign the disease outcome for each individual. Clinical expert adjudication was performed on 279 individuals to serve as the gold-standard label for algorithm validation.

The Michigan Genomics Initiative (MGI) is an EHR-linked biobank hosted at the University of Michigan with genotype data linked to EHR information to facilitate biomedical research. With both genetic and clinical data available for all individuals in MM-MGI, we were able to validate the prediction models using genetic and/or clinical information. The MM-MGI cohort used in this study is from data freeze 4 (release date: July 2021)[2].

The study cohorts were subset to individuals who self-reported as European American in the MM-HF and MM-MGI cohorts, to avoid having reduced performance of genetic predictors in non-white ancestries thereby biasing the model evaluation towards favoring clinical predictors. The MM-HF and MM-MGI cohorts were comprised of 90% and 86% European American individuals, respectively.

We refer to MM-PCP cohort as the code embedding derivation set, MM-HF cohort excluding individuals in MM-MGI cohort as ClinRS weights derivation set, and the intersection of MM-MGI and MM-HF cohort as model validation set. The model validation set has no overlap with the code embedding and ClinRS weights derivation sets (Supplementary Figure 1). First, the code embedding derivation set was used to learn EHR code patterns and build medical code embeddings for downstream analysis. Patients with a rich medical history and active records within the system were included for code co-occurrence pattern learning in the code embedding derivation set. Next, the labels curated in the MM-HF cohort served as the outcome in the ClinRS weights derivation set to obtain the weights to calculate ClinRS for heart failure cases prediction. The ClinRS weights derivation set consisted of 7,120 individuals from MM-HF and excluded those from the MM-MGI. Last, the model validation set (independent from ClinRS weights derivation set) was used to assess the prediction ability of PRS and ClinRS. The model validation set included 20,279 participants, who were drawn from the overlapping populations of the MM-MGI and MM-HF cohorts. All patients in the model validation set were assigned a label for heart failure outcome using a phenotyping, fully genotyped to calculate PRS, and had EHR data available to generate ClinRS (Supplementary Figure 1).

### Calculate Clinical Risk Score (ClinRS)

#### Extraction of medical code embeddings using NLP

The first step to summarizing the EHR data using NLP was to convert a patient's EHR medical codes from all healthcare encounters to paragraphs, then concatenate the patient's paragraphs of medical codes to create an article. After converting EHR data to an article, we were able to derive the co-occurrence patterns of each pair of medical codes. We extracted the semantic meaning of each code into numeric vector representations (medical code embeddings) that captured the clinically relevant information of each code. See supplementary materials - curating medical code embedding section in below for detailed explanation on the NLP approach to generate vector representation of medical codes.

#### Evaluation of NLP derived medical code embeddings and parameter tuning

The algorithm for obtaining the medical code embeddings as described above has two key tuning parameters: time window *t* and embedding dimension *d* (i.e., the number of features/ elements in a code embedding). The principle used in parameter tuning is to optimize the clinical meaningfulness of the medical code embedding. The code embeddings should capture similarity of the codes and thus be able to identify whether two specific codes describe the same overall medical concept (i.e., grouping of ICD codes).

To select the optimal time window *t* and embedding dimension *d* for the medical code embeddings, we developed a set of true labels for ICD code grouping using an expert curated ontology named phenome-wide association study code (phecode)[3]. Next, we evaluated whether code pairs that are mapped to the same phecode have larger cosine similarity (i.e. the cosine value of the angle between the corresponding medical code embedding vector pairs) than randomly selected pairs. The cosine similarity is a distance metric measuring how close the two codes are alike in terms of their concepts and meanings. It ranges from -1 to 1, with high cosine values representing that the selected pair of two codes have more similar semantic meaning and utilization context. These evaluations aid in the search for the most ‘clinically meaningful’ yet efficient version of medical code embedding with the smallest necessary dimension.

In summary, our goal was to find the optimal time window and embedding dimension that result in medical code embeddings that accurately capture the relationships between codes and represent the most clinically meaningful grouping of ICD codes.

In this analysis, phecodes are rolled up to the integer level[4]. For instance, ICD-9 code 428.2 (systolic heart failure) and 428.3 (diastolic heart failure) are mapped to the phecode 428.3 (heart failure with reduced EF) and 428.4 (heart failure with preserved EF), respectively. These two codes are then rolled up into to the same phecode group of 428. Moreover, both ICD-9 and ICD-10 codes can be mapped to the same phecode. For example, ICD-9 code 428.1 (left heart failure) and ICD-10 code I50.1 (left ventricular failure) are both mapped to phecode 428.2 (heart failure) and further rolled up to the integer 428.

To search for the most clinically meaningful medical code embeddings, we performed a classification task using phecode label and cosine scores. The classification label was the binary indicator of whether the two codes shared the same phecode. The classification score was the cosine distance score calculated between vector representations for two codes. This classification task showed whether a pair of codes mapped to the same phecode have a higher cosine similarity (similar semantic representations). The classification results were evaluated using Area Under the Receiver Operating Characteristics (AUC). To distinguish the AUC used in the subsequent evaluation of the heart failure prediction model, we refer to the AUC aiding grid search for optimal NLP derived medical code embeddings based on existing clinical concept ontology as concept-AUC. Concept-AUC is used throughout the remainder of this article to assess whether the medical code embeddings derived from NLP is clinically meaningful, in the sense that it can aid identifying whether arbitrary pairs of codes are describing the same concept or belonging to the same general group. The combination of time window *t* and embedding dimension *d* that achieved the highest concept-AUC was selected, the corresponding code embeddings were generated accordingly.

In the grid search for optimal time windows *t* and embedding dimension *d* combination, cosine similarity for 430,579,185 pairs of codes among 29,346 unique codes was calculated for each time window and embedding dimension combination. Ten *t* time windows (1, 2, 7, 10, 14, 20, 30, 40, 50, and 60 days) and twelve *d* embedding dimensions (10, 30, 50, 100, 150, 200, 250, 300, 350, 400, 450, and 500) were evaluated. This results in a total of 120 concept-AUC calculated to evaluate the clinical applicability of NLP-derived code concepts from EHR data.

**Calculation of patient-level latent phenotypes**

To create latent phenotypes for each patient, we used the medical code embeddings derived from the MM-PCP cohort curated from the previous step and applied this information to the diagnosis codes documented in the medical records of patients in the MM-HF cohort. Specifically, we summed up medical code embeddings corresponding to all codes present within a patient's medical record.

In detail, we took the product of the patient-level EHR record *D*, a dataset recorded whether patients had the diagnosis code in the past, and code embedding *C*, a semantic vector representation of the EHR codes. *D* is a n by p matrix, where n is the number of patients and p is the number of unique diagnosis codes. *C* is a p by k matrix, where p is the number of unique diagnosis codes and k is the embedding dimension selected from the code embedding curation – parameter tuning step. The final product of *D* and *C* will be the patient-level latent phenotypes with dimension of n by k. See Supplementary Figure 7 for illustration. These latent phenotypes summarize the information of a patient's medical diagnosis history.

**Time point specific latent phenotypes**

We sought to evaluate how far in advance we could predict heart failure and avoid label leakage. The rationale of avoiding label leakage is to not use nonexistent information in the prediction period to predict outcome, which could lead to overestimating the model performance. For example, we would like to avoid using the disease treatment or procedure information that is only available after disease diagnosis. To do this, we removed all ICD codes a year prior to the heart failure diagnosis date, then calculated the latent phenotypes. We repeated this procedure by increasing the exclusion time for ICD codes, starting from two years prior to disease diagnosis and then increasing the exclusion time in one-year increments up to ten years prior to disease diagnosis. This resulted in the generation of ten sets of latent phenotypes, each with a different cutoff time for medical history removal. Patients with no medical history recorded within the healthcare system prior to the cutoff time point were excluded from the analysis. See Supplementary Table 1 for sample size in each time point.

**Supervised training for ClinRS using LASSO**

To summarize the multi-dimensional patient-level latent phenotypes into a single risk score, we applied the Least Absolute Shrinkage and Selection Operator (LASSO) for feature selection with 10-fold validation for shrinkage parameter tuning[5]. The LASSO leverages the L1 penalty on the regression coefficients to eliminate non-important variables, avoid overfitting, and achieve better prediction. Next, the coefficients yielded from the LASSO model were used as weights (effect sizes) to calculate a weighted sum of patients’ clinical risk. In the ClinRS weights derivation set (individuals in MM-HF excluding MM-MGI), the patients' latent phenotypes were calculated using EHR records one year prior to heart failure diagnosis (Supplementary Figure 1). The heart failure outcome was regressed on 350 latent phenotypes and adjusted for age, sex, and healthcare utilization using logistic regression with L1 regularization. Three patient characteristics known to be strong predictors of the outcome (age, sex, and healthcare utilization) were forced in the model with no shrinkage. Patients' healthcare utilizations were summarized by the number of months of encounters recorded in the EHR.

**Calculate ClinRS for patients in model validation set**

To validate the prediction accuracy of ClinRS, we applied the ClinRS weights obtained from the ClinRS weights derivation set to an independent model validation set to summarize the entire EHR diagnosis records into one score (Supplementary Figure 1). The score was further used in the heart failure prediction model to predict patients' disease outcome in the future. For each participant in the model validation set, ten ClinRS were calculated using time point specific latent phenotypes from one year up to ten years prior to disease diagnosis. Next, we performed inverse normalization to convert the ClinRS score into standard normal distribution.

**Curating medical code embedding**

The medical code embeddings were obtained through an adapted NLP method that learned vector representations of ICD codes based on their co-occurrence patterns in the EHR[6]. More specifically, the medical code embeddings were extracted by performing truncated singular value decomposition (SVD) on the shifted positive pointwise mutual information (SPPMI) matrix, which is derived from codes' co-occurrence matrix. The pipeline we developed to extract medical code embedding was based on Hong *et al.*[7] and it is publicly available at https://github.com/The-Shi-Lab/CodeEmbedding.

### Co-occurrence Matrix

A co-occurrence matrix is defined with a selected time window *t*, within which the co-occurrence instances of codes are counted. Since there are 29,346 codes, the dimension of the co-occurrence matrix is 29,346-by-29,346, with each entry counting the number of co-occurrence instances in the EHR between the corresponding pair of codes. By this definition, the co-occurrence matrix is a symmetric matrix. Assuming that the selected time window $t$ is co-occurred within 7 days, for each code (which we denote by $C$) and each patient, we first identify the dates when the code was assigned to the patient. Then, for each of these identified dates, we scan the EHR of the patient within the day and the following 6 days; each code assignment found is counted as an instance of co-occurrence with code $C$. In such a fashion, the co-occurrence matrix is obtained by aggregating the co-occurrence instances over all patients and all codes.

### Calculation of medical code embedding

The medical code embeddings were obtained through dimension reduction of the SPPMI matrix, which is derived from the co-occurrence matrix, which we denote by $CC$. Specifically, the SPPMI matrix share the size of $CC$ which is 29,346-by-29,346 and for each code pair $C_1, C_2$,

$$SPPMI(C_1, C_2) = max\{log\frac{CC(C_1,C_2)}{CC(C_1,\cdot)CC(C_2,\cdot)} - log(k), 0\}$$

where $CC(C_1,\cdot)$ represents the row sum of $CC$ on the row corresponding to $C_1$. The tuning parameter, negative sample $k$ was set to 10 based on results shown in previous studies[8–10]. Given a SPPMI matrix and a desired semantic vector representation (SEV) dimension $d$, the SEVs are obtained through the truncated singular value decomposition of the SPPMI matrix, which we denote by $U_d diag(\sigma_1, \cdots, \sigma_d) {U_d}^T$, where $\sigma_1 \geq \sigma_2 \geq \cdots \geq \sigma_d$ are the $d$ largest singular values of the SPPMI matrix. Specifically, the $d$ SEVs are the columns of $U_d diag(\sqrt{\sigma_1}, \cdots, \sqrt{\sigma_d})$, which are all vectors with 29,346 entries (one for each ICD code).

**Sensitivity analysis removing circulatory system diagnosis codes**

To further verify the validity of ClinRS, additional analyses were conducted to examine the robustness of the co-occurrence patterns captured by the unsupervised NLP algorithm. We created a ClinRS without circulatory system information (ClinRS-NoCirc) by excluding ICD diagnosis codes belonging to ICD-9 Seventh Chapter (390-459) and ICD-10 Chapter IX (I00-I99): Diseases of the Circulatory System. The ClinRS without circulatory system was further used in model prediction to evaluate the ability of the proposed method. The goal of the sensitivity analysis was to predict disease outcome (heart failure) without directly associated diagnosis information (circulatory system diagnosis codes). We excluded 1,340 circulatory system diagnosis codes (459 from ICD-9 and 881 from ICD-10) and used the rest of the 28,006 codes to create patient-level latent phenotypes, and applied the newly derived latent phenotypes with ClinRS weights derived previously to generate ClinRS-NoCirc. We demonstrated that using pre-trained co-occurrence patterns from an independent dataset could be valuable for disease prediction and the co-occurrence patterns aided capturing disease risks through indirect associations.

### RESULTS

#### NLP extracted medical code embeddings are clinically meaningful

We discovered two main findings: 1) smaller time window size *t* and 2) inclusion of more features *d* in a code embedding yielded higher accuracy on identifying code pairs in the same phecode group. Supplementary Figure 2 showed that holding constant embedding dimension *d* while varying time window size *t*, the highest concept-AUC was consistently found from co-occurrence matrices constructed based on codes that appeared on the same day (within 1 day). The accuracy attenuated linearly when the window size increased. For example, concept-AUC calculated from embedding dimension of 350 was the highest for codes co-occurred on the same day (1 day) with concept-AUC of 0.78, decreased to 0.76 for codes co-occurred within 1 week (7 days), and dropped to the lowest of 0.73 for codes that co-occurred within 2 months (60 days).

Next, we evaluated the concept-AUC variation across different numbers of embedding dimension *d* in a code embedding. In general, the higher the embedding dimension *d*, the higher the concept-AUC was observed. The optimal embedding dimension found in this study using Michigan Medicine EHR data was $d = 350$ (Supplementary Figure 2). The medical code embeddings generated from time window $t = 1$ day with embedding dimension $d = 350$ yielded the highest concept-AUC of 0.78 in this study (Supplementary Figure 2).

#### Sensitivity analysis on removing circulatory system diagnosis code

To examine the robustness of ClinRS and address concerns regarding overfitting, we conducted a sensitivity analysis by removing all circulatory system diagnosis codes to create ClinRS-NoCirc. In Supplementary Figure 6, we presented the model performances of using ClinRS-NoCirc as the clinical risk predictor and compared to the ClinRS model. The results were largely consistent with and without removing circulatory system codes, which demonstrated the success of our efforts to build a risk score that leveraged the high-dimensional EHR records and summarized underlying patterns to reveal disease associations. Notably, the models using ClinRS-NoCirc for predicting future heart failure events yielded significantly higher accuracy than baseline models, up to six years in advance of disease diagnosis. We observed an AUC of 0.77 (0.75-0.80) from ClinRS-NoCirc model at six years prior to disease diagnosis, which was significantly higher than baseline model at six years in advance of heart failure diagnosis (AUC: 0.72 [0.69-0.74]) (see Supplementary Figure 6 and Supplementary Table 1). Although the results derived from ClinRS-NoCirc could not predict the outcome as many years in advance as the ClinRS model, the additive power of integrating genetic and clinical information in disease risk prediction remains evident through ClinRS-NoCirc. By including both PRS and ClinRS-NoCirc in the heart failure prediction model, we were still able to distinguish patients with high risk of heart failure a decade in advance of the disease diagnosis. The heart failure prediction model with PRS and ClinRS-NoCirs predictors showed a significantly higher AUC of 0.78 (0.76-0.81) at ten years prior to heart failure diagnosis, compared to the baseline model with AUC of 0.72 (0.69-0.75).

**TABLES and FIGURES**

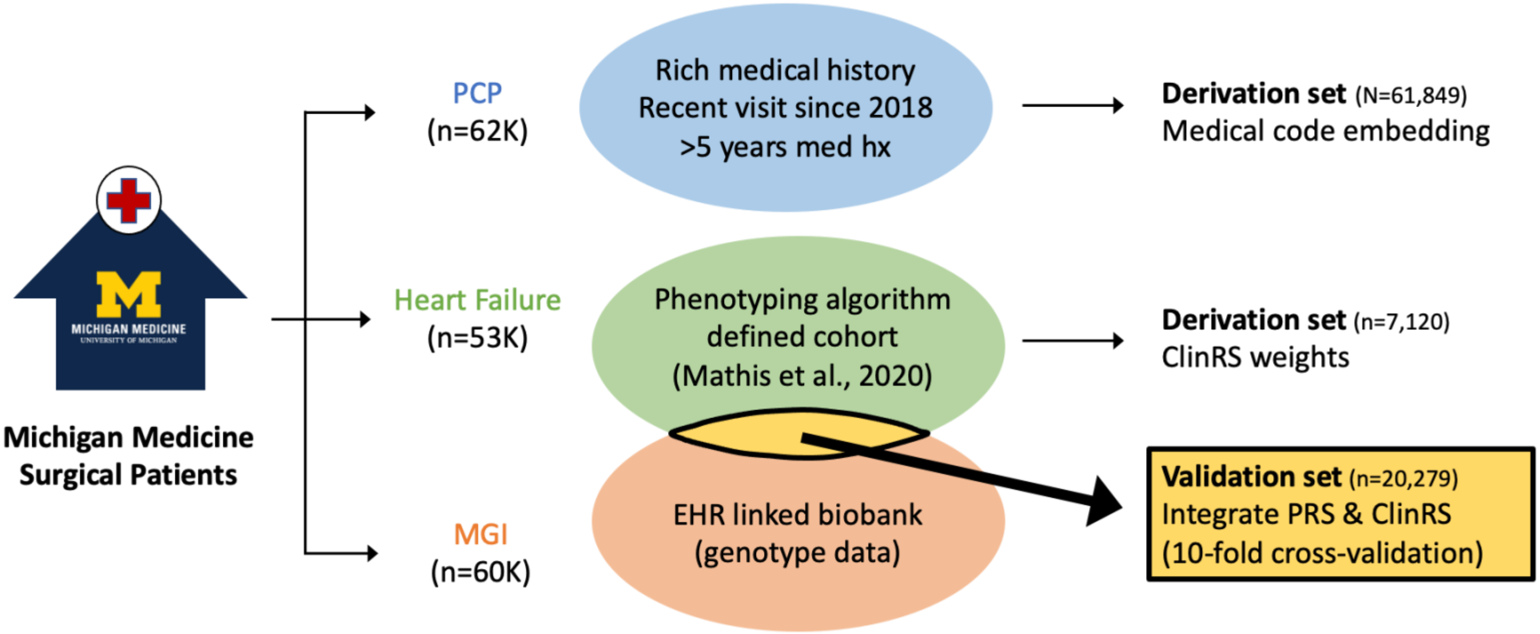

**Supplementary Figure 1.** Three cohorts within Michigan Medicine (MM) were used in this analysis: i) Primary Care Provider (MM-PCP), ii) Heart Failure (MM-HF), and iii) Michigan Genomics Initiative (MM-MGI). MM-PCP cohort with 61,849 individuals was used to build medical code embeddings. Subset of MM-HF (N=7,120), participants of European descent and not in MM-MGI, was used to derive the weights (effect sizes) of clinical risk score (ClinRS). Subset of MM-MGI (N=20,279), patients fully genotyped and disease outcome was predefined using Mathis et al. phenotyping algorithm[1] in MM-HF, was used to validate heart failure prediction accuracy using polygenic risk score and clinical risk score.

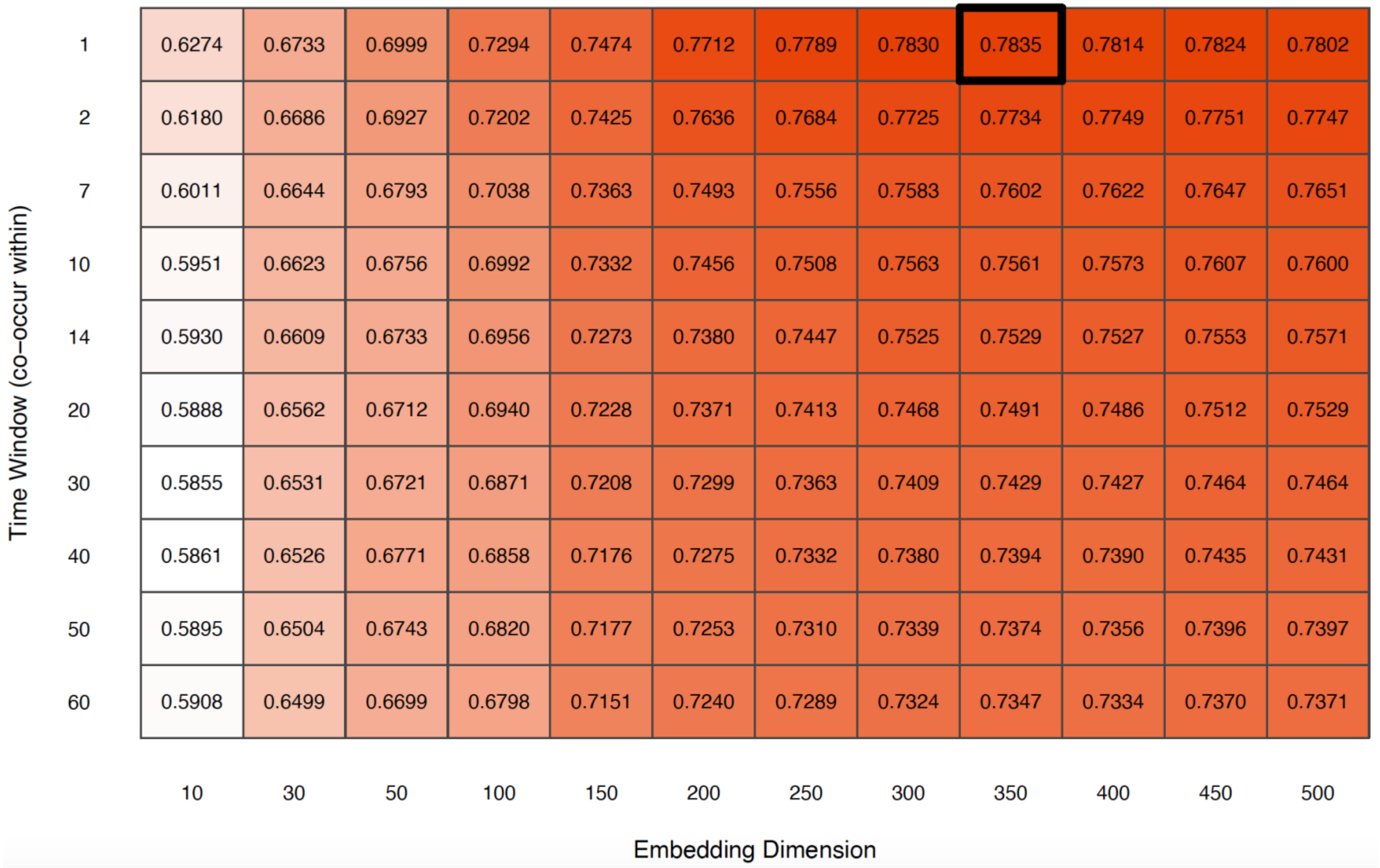

**Supplementary Figure 2.** Heatmap of concept-AUC across medical code embeddings derived from using 10 time windows and 12 embedding dimensions to summarize a medical code. Concept Area Under the Receiver Operating Characteristics (concept-AUC) summarized how well medical code embeddings generated from the adapted natural language (NLP) processing method capture the clinical meaning of each code. Medical code embedding built on code co-occurred within 1 day with embedding dimension of 350 yielded the highest concept-AUC.

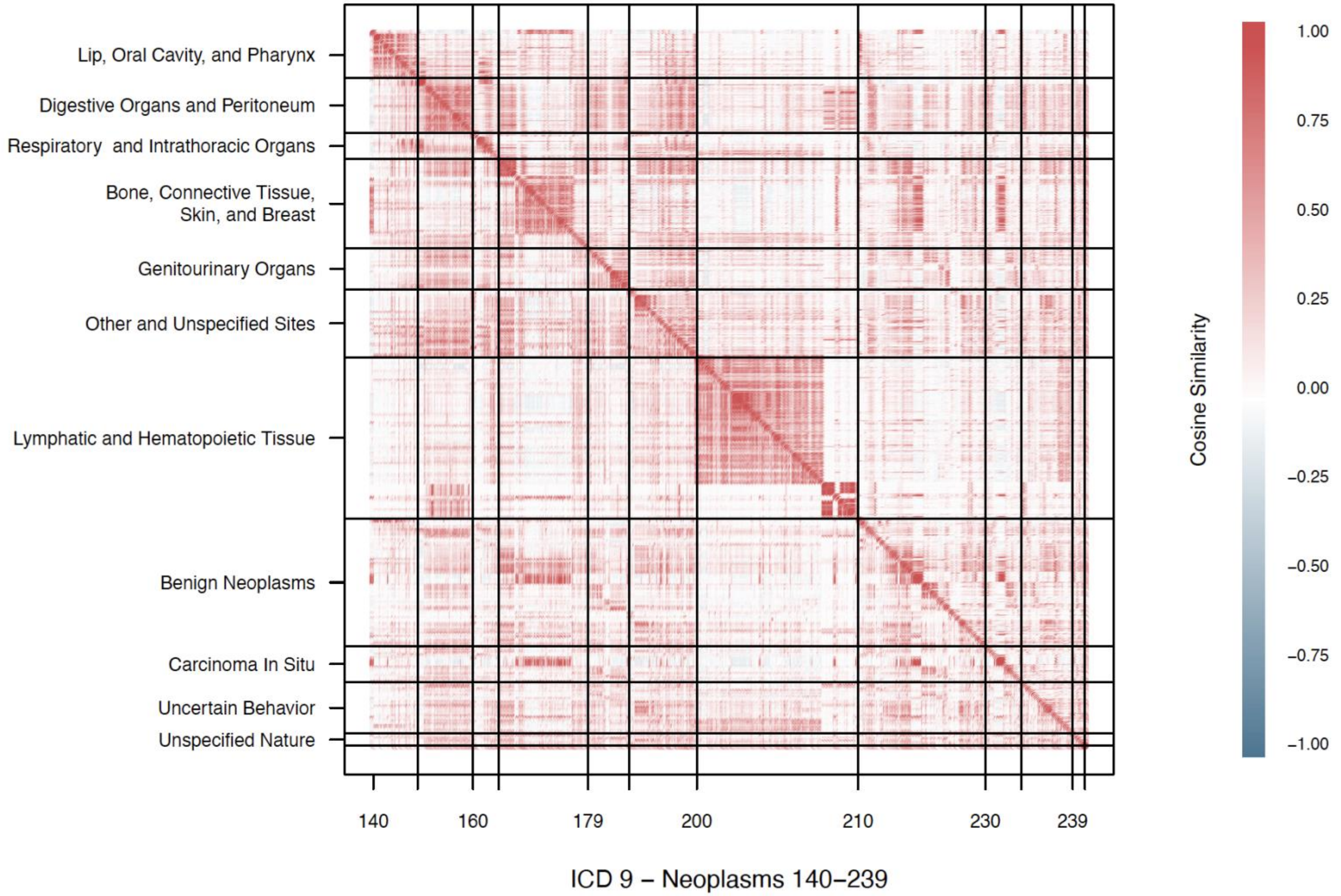

**Supplementary Figure 3.** Heatmap of cosine similarity score between a pair of codes within ICD-9 140 to 239 (Neoplasms) and sorted by its order. Every dot in this plot represents a pair of codes and its cosine similarity score, with the darker the red representing the closer the distance (more similar) between these 2 codes.

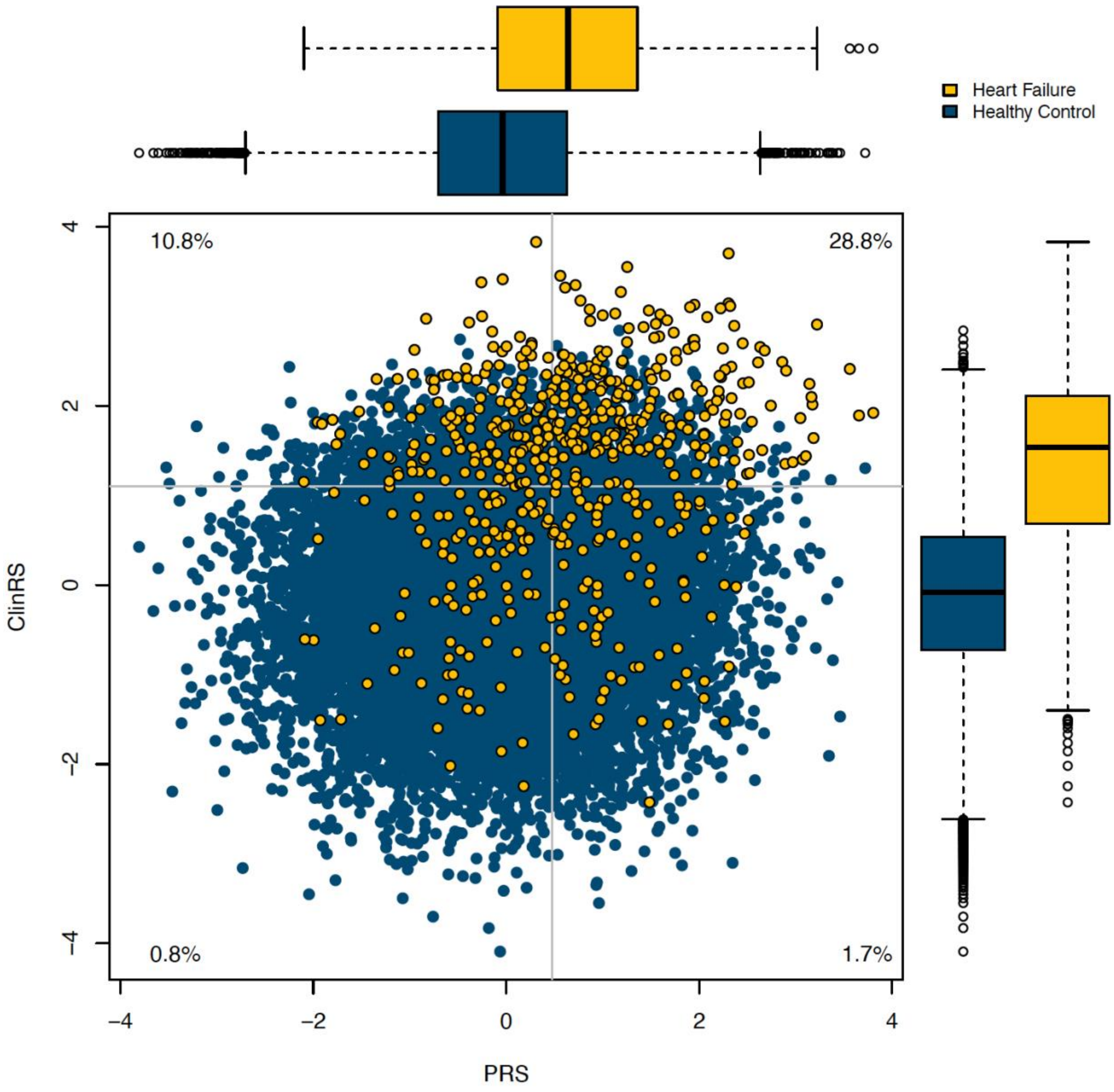

**Supplementary Figure 4.** Scatter plot and boxplot of patients' polygenic risk score (PRS) and clinical risk score (ClinRS) at one year prior to heart failure diagnosis, colored by disease status. Dotted gray lines indicate the cutoff of high and low risk of corresponding risk predictors. Percentage in each quadrant indicates the percentage of heart failure cases among patients classified in the corresponding risk group.

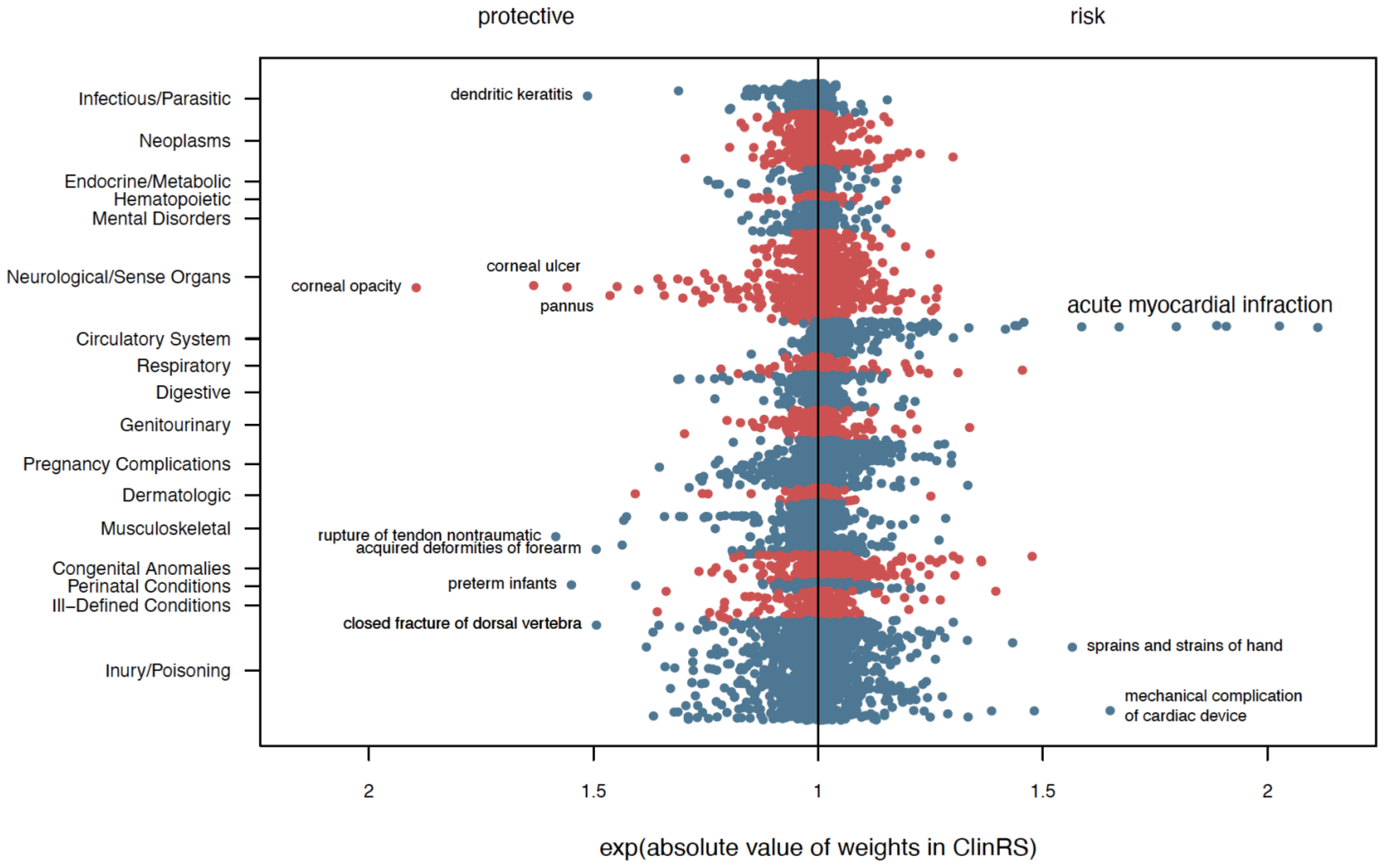

**Supplementary Figure 5.** Manhattan plot of clinical risk score (ClinRS) weights for each ICD-9 diagnosis code by disease class. X-axis indicates the exponential of the absolute weights in ClinRS. The left panel showed the weights of the protective (negative weights; decreased risk) factor and the right panel showed the weights of the risk (positive weights; increased risk) factor.

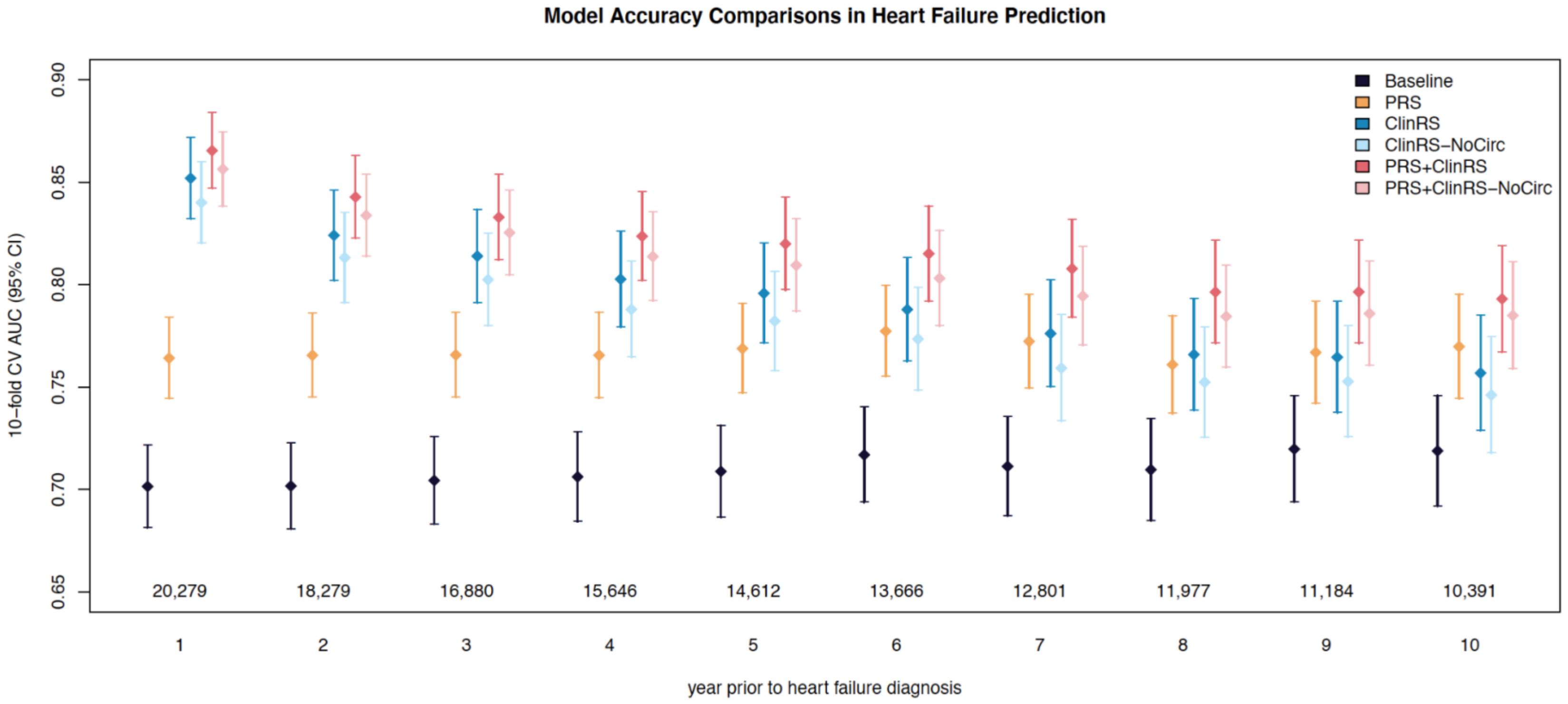

**Supplementary Figure 6.** Forest plot comparing models accuracy of predicting heart failure at one to ten years prior to disease diagnosis in the sensitivity analysis. Six models were compared with each time point: baseline (age and sex), PRS (polygenic risk score), ClinRS (clinical risk score), ClinRS-NoCirc, PRS+ClinRS, and PRS+ClinRS-NoCirc. ClinRS-NoCirc was calculated by removing circulatory system diagnosis code in patients' medical records to validate the validity of ClinRS generated using the adapted natural language processing method. Numbers at the bottom of the plot indicate the sample size for each time point. Results showed that ClinRS-NoCirc can predict heart failure outcomes six years in advance, shorter than using ClinRS as a predictor. Adding both PRS and ClinRS-NoCirc in the model, the model accuracy is comparable to PRS+ClinRS model, which predicts disease ten years in advance.

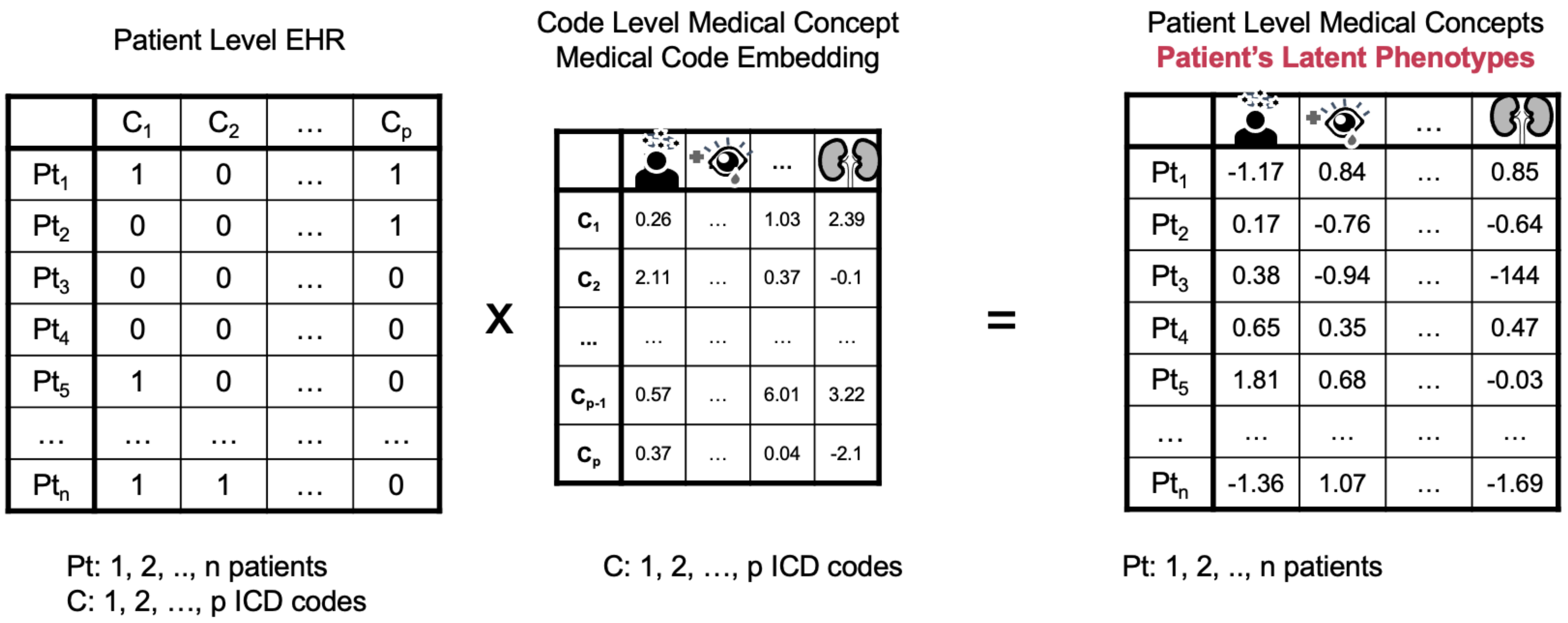

**Supplementary Figure 7.** Illustration of creating latent phenotype from individual level electronic health records.

**Supplementary Table 1.** Sample size of heart failure cases and controls included in analysis for one to ten years prior to disease diagnosis.

| Year | Sample Size | | 10-fold Cross-Validated AUC | | | | | | |
|---|---|---|---|---|---|---|---|---|---|
| | cases | controls | baseline | ARIC | PRS | ClinRS | PRS+ClinRS | ClinRS-noCirc | PRS+ ClinRS-noCirc |
| 1 | 576 | 19,703 | 0.70 (0.68-0.72) | 0.81 (0.79-0.83) | 0.76 (0.74-0.78) | 0.85 (0.83-0.87) | 0.86 (0.85-0.88) | 0.84 (0.82-0.86) | 0.86 (0.84-0.88) |
| 2 | 539 | 17,758 | 0.70 (0.68-0.72) | 0.81 (0.79-0.82) | 0.76 (0.74-0.79) | 0.82 (0.80-0.85) | 0.84 (0.82-0.86) | 0.81 (0.79-0.84) | 0.83 (0.81-0.85) |
| 3 | 515 | 16,365 | 0.70 (0.68-0.73) | 0.79 (0.77-0.81) | 0.77 (0.74-0.79) | 0.81 (0.79-0.84) | 0.83 (0.81-0.85) | 0.80 (0.78-0.82) | 0.82 (0.80-0.85) |
| 4 | 494 | 15,152 | 0.71 (0.68-0.73) | 0.79 (0.77-0.81) | 0.76 (0.74-0.79) | 0.80 (0.78-0.83) | 0.82 (0.80-0.84) | 0.79 (0.76-0.81) | 0.81 (0.79-0.84) |
| 5 | 459 | 14,153 | 0.71 (0.69-0.73) | 0.77 (0.75-0.79) | 0.77 (0.75-0.79) | 0.80 (0.77-0.82) | 0.82 (0.80-0.84) | 0.78 (0.76-0.81) | 0.81 (0.79-0.83) |
| 6 | 427 | 13,239 | 0.72 (0.69-0.74) | 0.78 (0.76-0.80) | 0.78 (0.76-0.80) | 0.79 (0.76-0.81) | 0.81 (0.79-0.84) | 0.77 (0.75-0.80) | 0.80 (0.78-0.83) |
| 7 | 407 | 12,394 | 0.71 (0.69-0.74) | 0.77 (0.75-0.80) | 0.77 (0.75-0.80) | 0.78 (0.75-0.80) | 0.81 (0.78-0.83) | 0.76 (0.73-0.78) | 0.79 (0.77-0.82) |
| 8 | 376 | 11,601 | 0.71 (0.69-0.74) | 0.76 (0.74-0.79) | 0.76 (0.74-0.78) | 0.77 (0.74-0.79) | 0.80 (0.77-0.82) | 0.75 (0.73-0.78) | 0.78 (0.76-0.81) |
| 9 | 353 | 10,831 | 0.72 (0.69-0.75) | 0.77 (0.74-0.80) | 0.77 (0.74-0.79) | 0.76 (0.74-0.79) | 0.80 (0.77-0.82) | 0.75 (0.73-0.78) | 0.79 (0.76-0.81) |
| 10 | 332 | 10,059 | 0.72 (0.69-0.75) | 0.77 (0.74-0.79) | 0.77 (0.74-0.80) | 0.76 (0.73-0.78) | 0.79 (0.77-0.82) | 0.75 (0.72-0.77) | 0.78 (0.76-0.81) |

Ten-fold cross-validated Area Under the Receiver Operating Characteristics (AUC) of six models predicting heart failure outcome across 10 time points. Model performances were calculated for baseline (age and sex) model and 5 models with risk score(s) added: i) polygenic risk score (PRS), ii) clinical risk score (ClinRS), iii) PRS+ClinRS, iv) clinical risk score calculated without circulatory system diagnosis code (ClinRS-NoCirc), and v) PRS+ClinRS-NoCirc.

**Supplementary Table 2.** Top 20 protective and risk factors yielded from clinical risk score (ClinRS).

| Protective Factors | | | Risk Factors | | |
|---|---|---|---|---|---|
| **ICD code** | **ClinRS weight** | **Diagnosis** | **ICD code** | **ClinRS weight** | **Diagnosis** |
| 371.03 | -0.6391 | Opacity, central cornea | 410.91 | 0.7476 | AMI NOS, initial |
| 370.03 | -0.4905 | Ulcer, central corneal | 410.21 | 0.7063 | AMI, inferolateral wall, initial |
| 727.63 | -0.4600 | Rupture, hand/wrist extensor tendon | 410.41 | 0.6463 | AMI, inferior wall, initial |
| 370.63 | -0.4441 | Vascularization, deep corneal | 410.01 | 0.6400 | AMI, anterolateral wall, initial |
| 765.14 | -0.4378 | Preterm infant NEC, 1000-1249 gram | 410.51 | 0.5861 | AMI, lateral wall, initial |
| 54.42 | -0.4144 | Herpes simplex dendritic keratitis | 410.71 | 0.5127 | AMI, subendocardial, initial |
| 736.09 | -0.4015 | Deformity, acquired, forearm NEC | 996.01 | 0.5007 | Malfunction, cardiac pacemaker |
| 806.25 | -0.4011 | Fx T7-T12 clsd w/spinal cd inj NOS | 410.61 | 0.4616 | True posterior wall, initial |
| 374.23 | -0.3808 | Lagophthalmos, cicatricial | 842.19 | 0.4483 | Sprain/strain, hand NEC |
| 370.35 | -0.3697 | Keratoconjunctivitis, neurotrophic | 996.04 | 0.3927 | Complications d/t AICD |
| 732.7 | -0.3621 | Osteochondritis dissecans | 743.37 | 0.3894 | Ectopic lens, congenital |
| 718.84 | -0.3596 | Drngmnt, oth joint NEC, hand | 396 | 0.3768 | Stenosis, mitral and aortic valves |
| 717.89 | -0.3554 | Disruption, internal, knee NEC | 512.8 | 0.3746 | Pneumothorax, spontaneous NEC |
| 695.14 | -0.3416 | SJS toxic epidermal necrolysis synd | 410.11 | 0.3674 | AMI, anterior wall, initial |
| 765.25 | -0.3407 | Gestation completed 29-30 weeks | 410.02 | 0.3631 | AMI, anterolateral wall, subsequent |
| 371.61 | -0.3364 | Keratoconus, stable | 835.03 | 0.3596 | Dsloc, anterior hip NEC, closed |
| 842.12 | -0.3239 | Sprain/strain, metacarpophalangeal | 414.2 | 0.3483 | Chrn total occlusion coronary arter |
| 813.54 | -0.313 | Fx lower radius w/ulna, open | 780.32 | 0.3331 | Symp, convulsions, febrile complex |
| 997.4 | -0.3124 | Complications, digestive system | 996.09 | 0.3264 | Malfunction, cardiac dev/graft NEC |
| 793.1 | -0.3062 | AbFnd, rdlog, lung field | 746.86 | 0.3102 | Block, heart, congenital |
